# Clinical performance of proenkephalin A 119-159 for the early diagnosis of acute kidney injury in patients with sepsis or septic shock

**DOI:** 10.1101/2024.10.11.24315291

**Authors:** Janin Schulte, François Dépret, Oliver Hartmann, Peter Pickkers, Pierre-François Laterre, Florian Uhle, AdrenOSS-1 study investigators

## Abstract

**Background:** Sepsis-associated acute kidney injury (AKI) is a severe condition associated with unfavorable outcomes in critically ill patients, not least because of its delayed diagnosis and management due to limitations in the current standard of care. We aimed to evaluate the performance of sphingotest® penKid® for the early diagnosis of AKI in patients with sepsis or septic shock.

**Methods:** Plasma proenkephalin A 119–159 (penKid) was measured in 120 healthy subjects and 529 critically ill sepsis patients. Subgroup analyses were performed for patients with and without a history of heart failure or hypertension. The clinical performance for the diagnosis of AKI within 48 hours (AKI_48h_) was calculated using the upper limit of the penKid reference range. To improve clinical decision-making, interpretation bands and likelihood ratios for optimized rule-in and rule-out performance were established.

**Results:** Of the 529 patients with sepsis or septic shock, 328 were male (62%), and the median age was 66 (interquartile range 56–75) years. Two hundred thirty-four (44%) patients were diagnosed with AKI_48h_, and those patients presented increased penKid levels on intensive care unit admission compared with patients without AKI_48h_, with an area under the curve of 0.87 (95% confidence interval [CI] 0.84–0.90, p<0.0001). A penKid cut-off of 89 pmol/L, corresponding to the 97.5^th^ percentile in the healthy reference population, resulted in 72% sensitivity (95% CI 66–77%), 83% specificity (95% CI 78–87%), 77% positive predictive value (95% CI 71–82%), 79% negative predictive value (95% CI 74–83%), a positive likelihood ratio of 4.2, and a negative likelihood ratio of 0.34 to diagnose AKI_48h_. An improved performance was obtained when patients with a history of chronic heart failure and/or hypertension were excluded. PenKid cutoff values of 54 pmol/L (>92% sensitivity) and 105 pmol/L (>92% specificity) were derived to establish actionable interpretation bands for diagnostic rule-in and rule-out.

**Conclusions:** The sphingotest® penKid® assay can facilitate an early diagnosis of AKI up to 48 hours before the KDIGO criteria are met. Based on the present results, the assay was registered as an aid in the early diagnosis of AKI in patients with sepsis or septic shock.

## Background

Acute kidney injury (AKI) affects approximately 10–15% of patients admitted to the hospital (1). In critically ill patients, its incidence has been reported to exceed 50% (1, 2), which appears to be increasing (3). Sepsis, which is characterized by life-threatening organ dysfunction due to a dysregulated host response to infection (4), is associated with a greater risk of developing AKI (5), and the costs associated with AKI management impose a significant burden on healthcare systems worldwide (6). The negative consequences of AKI extend beyond the acute phase, with progression to acute kidney disease (no recovery from AKI after seven days) and chronic kidney disease (CKD) (entering CKD after recovery from AKI) (7, 8), an increased risk of cardiovascular complications, recurrent episodes of AKI, and long-term mortality (2, 9). The current definition of AKI was released in 2012 by the AKI work group of the Kidney Disease Improving Global Outcomes (KDIGO) organization and is based on an increase in serum creatinine and/or a decrease in urine output as surrogate markers of the glomerular filtration rate (GFR) (10). Serum creatinine is a breakdown product of muscle metabolism and is heavily affected by confounding factors such as volume status, muscle mass, and nutritional status, each individually causing differences in production (11). Serum creatinine reaches equilibrium between production and elimination with a time delay of hours to days, hampering both real-time reflection of the GFR in non-steady-state conditions and accurate AKI diagnosis (11, 12). Urine output may also be influenced by non-GFR-related factors such as hypovolemia and the use of diuretics, and is an imperfect marker lacking sensitivity and specificity (11, 12).

Circulating proenkephalin A 119–159 (penKid) is a stable peptide fragment cleaved during posttranslational maturation of the precursor preproenkephalin and is useful as a surrogate marker of enkephalins, endogenous opioids that bind predominantly to the delta opioid receptor. Apart from the high density of delta opioid receptors in the kidneys and the direct effects of enkephalins on natriuresis and diuresis (13, 14), proenkephalin is filtered in the glomerulus while no tubular or other excretion occurs and is therefore considered a functional biomarker for GFR (13). Various clinical studies have shown that penKid strongly correlates with GFR over the full range (15), facilitates early detection of AKI, and serves as a valuable aid for the prediction of liberation success from acute renal replacement therapy (RRT) (16–19). The performance of penKid in diagnosing AKI was recently summarized in a meta-analysis using data from patients with various critical conditions, such as sepsis, acute heart failure, and cardiac surgery (20).

In two clinical performance studies using biobanked plasma samples from healthy subjects and sepsis or septic shock patients, we aimed to evaluate the performance of sphingotest® penKid® as an aid in the early diagnosis of AKI.

## Methods

### Reference population

Ethylenediaminetetraacetic acid (EDTA) plasma samples, which were collected from self-reported healthy adult subjects between June 2016 and May 2021 and stored at −18 °C, were procured from a local biobank (in.vent Diagnostica GmbH). Healthy male or female subjects who were 18 years of age or older and willing to provide written informed consent were eligible for the study and completed a health questionnaire. Subjects were excluded if they were unable or unwilling to provide written informed consent; had a known pregnancy; had a current or recent (<10 years) history of cardiovascular disease; had diseases of the kidneys, lungs, vascular system, skin, gastrointestinal tract, liver, biliary tract, gallbladder, nervous system, epilepsy or autoimmune diseases; endocrine diseases; diseases of the hematopoietic system; rheumatic and/or immunological diseases; asthma; undefined skin eczema; gynecological diseases; or cancer, as well as infectious diseases at or within eight weeks before study enrollment. According to the Clinical and Laboratory Standards Institute (CLSI) approved guideline EP28-A3c, the minimum sample size of 120 subjects was used for adequate representation of females and males, as well as for reflecting adult age subgroups, including subjects between 18 and 39 years of age, subjects between 40 and 59 years of age, and subjects older than 60 years of age.

### AdrenOSS-1 study

The observational study Adrenomedullin and Outcome in Sepsis and Septic Shock-1 (AdrenOSS-1) was conducted between June 2015 and May 2016 at 24 centers in France, Belgium, the Netherlands, Italy and Germany and included patients above 18 years of age with a diagnosis of severe sepsis or septic shock on intensive care unit (ICU) admission or after transfer from another ICU within 24 hours (ClinicalTrials.gov identifier: NCT02393781) (21). Severe sepsis and septic shock were defined according to sepsis-2 (22). Following sepsis-3 definition the term “severe sepsis” has been rephrased to “sepsis” in the present article (4). Patients who were pregnant, in vegetative coma, or who participated in an interventional trial in the preceding month were excluded. The study was performed after local ethics approval was obtained and in compliance with good clinical practice and the Declaration of Helsinki. Patient data, including demographics, comorbidities, laboratory parameters, use of organ support, and severity scores (SOFA and acute and physiology and chronic health evaluation II [APACHE II]), were collected upon admission. Patients were followed up for a maximum of 90 days for recording of clinical endpoints such as mortality. Ethylenediaminetetraacetic acid plasma samples for penKid analysis were obtained on the day of ICU admission (median 13.0 hours, interquartile range [IQR] 4.5–18.5 hours) and stored at −80 °C until use.

Among the 585 patients in the AdrenOSS-1 study, 56 patients were excluded because of nonavailability of EDTA plasma for measurement of penKid or missing endpoint data that occurred because of discharge or death within 48 hours.

### Endpoint definition

Acute kidney injury diagnosis was defined in line with the practical approach of the KDIGO criteria (10), not using the urine output criterium but a change in serum creatinine ≥0.3 mg/dL (stage 1), serum creatinine 2.0–2.9 times baseline (stage 2), and serum creatinine ≥3.0 times baseline or the need for RRT within a maximum of 48 hours (AKI_48h_), i.e. early sepsis-associated AKI (23). The first serum creatinine value obtained after admission to the ICU served as the baseline reference value.

### Measurement of penKid

The measurement of penKid in EDTA plasma samples was performed in September 2021 by a certified service laboratory blinded to the clinical and demographic data of the subjects/patients and using a nonautomated microtiter plate (MTP)-based immunoluminometric assay (ILMA) sphingotest® penKid® (SphingoTec GmbH, Hennigsdorf, Germany), as described earlier (24). According to the instructions for use, the sphingotest® penKid® has a limit of detection and limit of quantitation of 29.9 pmol/L. The analyte penKid is stable in EDTA plasma for up to 48 hours at room temperature at 15–30 °C or refrigerated at 2–8 °C and for up to five freeze-thaw cycles. In addition, long-term stability was analyzed and revealed no significant difference in penKid levels in the clinical study samples after five years of storage at −80 °C.

### Statistical analysis

The 97.5^th^ percentile and 90% confidence interval (CI) for the distribution of penKid in the reference population was computed following the general procedures outlined in section 9 of the CLSI-approved guideline EP28-A3c.

To test for outliers, the outlier test ratio D/R was applied, where D is the absolute difference between the extreme observation and the next largest observation, and R is the range of all observations (25).

To evaluate the clinical performance of penKid as an aid in the early diagnosis of AKI, sensitivity and specificity, positive (PPV) and negative (NPV) predicted values, and the diagnostic odds ratio (OR) (all including 95% CI) were determined as performance indicators, using the 97.5^th^ percentiles of the distribution of penKid in the reference population as cut-off values, following the recommendations of MedTech Europe (Clinical evidence requirements for CE certification under the diagnostic regulation in the European Union, second edition). A 95% exact binomial CI for both sensitivity and specificity was computed. A test of the hypothesis of no association between penKid and the proportion of patients with a diagnosis of AKI was conducted via Fisher’s exact test, with a significance level of α = 0.05. The positive predictive value (PPV), NPV, positive likelihood ratio (LR+), negative likelihood ratio (LR-), and OR were computed accordingly. A minimum of 452 subjects was required to prove that both the sensitivity and specificity were above 60%, assuming that the true sensitivity and specificity were above 70%. Receiver operating characteristics (ROC) curves were constructed, and the area under the ROC curve (AUC) was calculated.

Patients were grouped into three different interpretation bands of AKI probability based on distinct penKid cutoffs at 92% sensitivity (rule-out) and 92% specificity (rule-in). The LR+ was calculated for the two upper interpretation bands, whereas the LR-was calculated for the lower interpretation band. The pretest probability corresponds to the prevalence of AKI in the AdrenOSS-1 study. The posttest probabilities were calculated as likelihood ratios of each band, i.e., true positives/(true positives+false positives).

Values are expressed as medians and IQR, or counts and percentages as appropriate. Group comparisons of continuous variables were performed via the Kruskal-Wallis test. Categorical data were compared via Pearson’s chi-square test.

For all the statistical analyses, R version 4.2.2 (http://www.r-project.org) or IBM SPSS Statistics Version 22 was used.

## Results

### Proenkephalin A 119–159 reference range

In the initial set of 120 healthy subjects, one subject (female, 54 years of age) with a penKid level of 158 pmol/L was considered an outlier according to statistical outlier testing. Therefore, this subject was excluded and replaced by a backup subject. Among the final 120 healthy subjects in the reference population, 65 (54%) were male. The median age was 51 (IQR 40– 59) years, with 28 subjects (23%) being 18–39 years, 63 subjects (53%) 40–59 years and 29 subjects (24%) 60 years of age or older. Concerning penKid levels, the population’s 97.5^th^ percentile was 89.3 pmol/L (90%CI 85.0–118.1 pmol/L). The median penKid level was 49.0 (IQR 39.5–60.9) pmol/L in all the subjects, with higher penKid levels in females than in males (54.0 [IQR 42.2–63.6] pmol/L vs. 46.1 [IQR 21.1–57.5] pmol/L, p=0.0308).

### Patient characteristics

The baseline characteristics and clinical outcomes of the 529 patients with sepsis or septic shock included in the AdrenOSS-1 study are summarized in Table 1. The patients had a median age of 66 (IQR 56–75) years, 328 (62%) patients were male, and 266 (50%) patients were diagnosed with septic shock on admission. The majority of patients were admitted for medical reasons (430 [81%] patients) rather than surgical reasons (99 patients [19%]). The primary site of infection was the lung (203 [38%] patients), followed by primary bloodstream (83 [16%] patients) and urinary tract infections (57 [11%] patients). The majority of patients had at least one cardiac (68%) or noncardiac (71%) comorbidity. The cohort presented a median SOFA score of 7 (IQR 5–10) points and a median APACHE II score of 16 (IQR 12–20) points. The majority of patients received vasopressors and/or inotropes (320 [61%] patients) and were on mechanical ventilation (321 [61%] patients) on admission. Forty-eight (9%) patients received RRT. The median ICU length of stay was 5.5 (IQR 2–10) days; 107 patients (20%) died within 28 days, and 144 patients (27%) died within 90 days after admission.

**Table 1:**
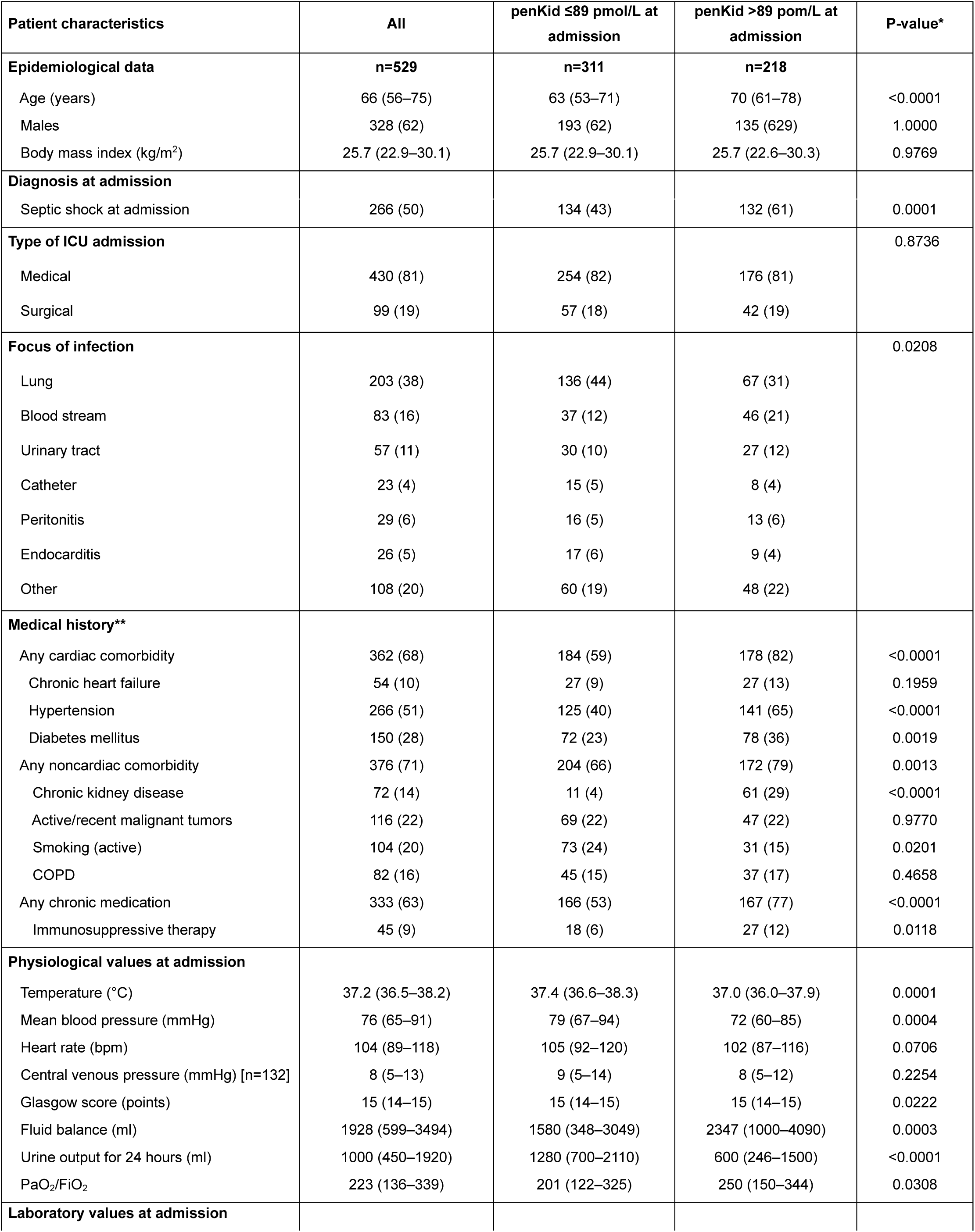
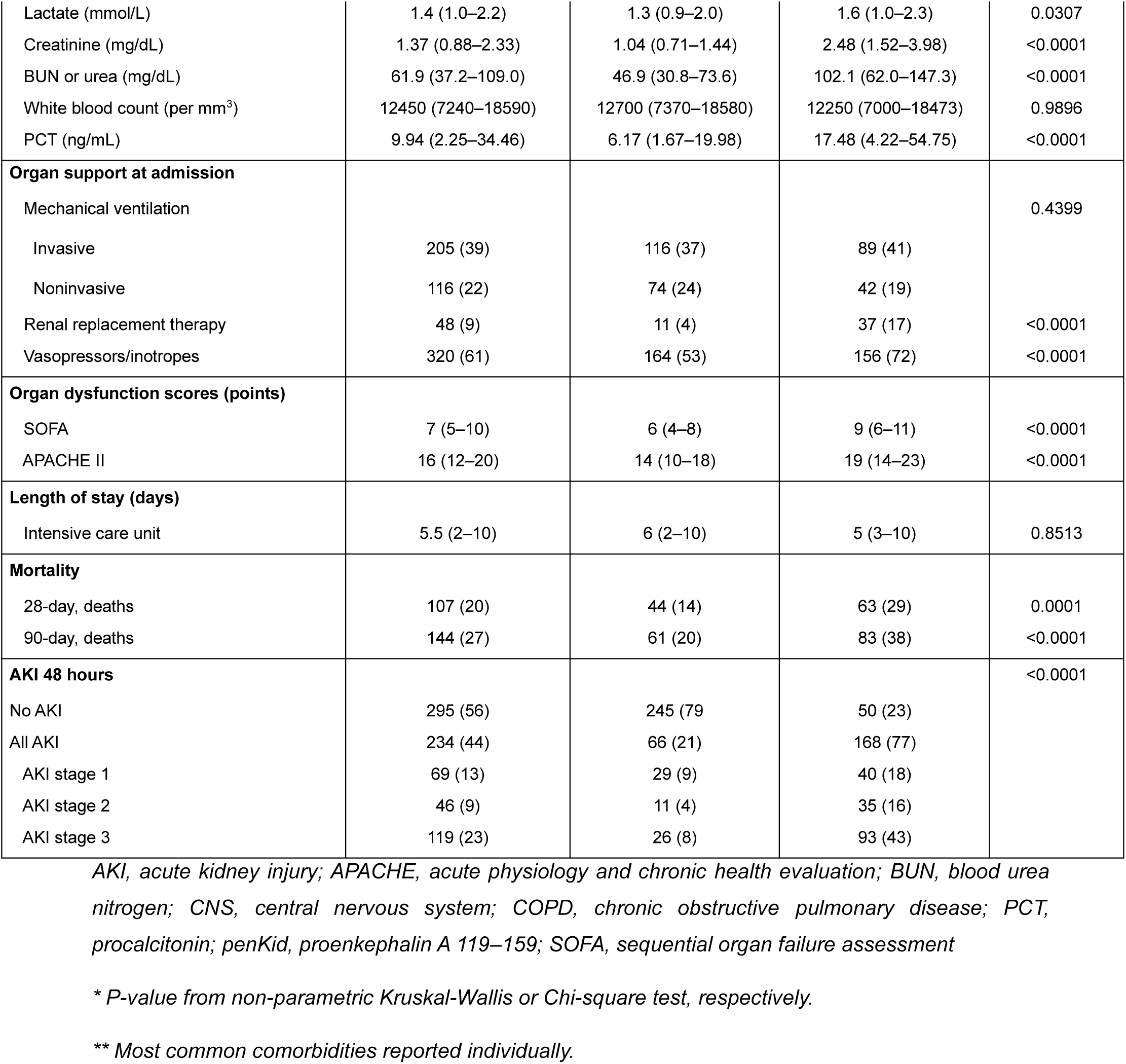
Patient characteristics and clinical outcomes.

Patients with a penKid equal or above to 89 pmol/L at ICU admission were older (median age 70 [IQR 61–78] vs. 63 [IQR 53–71] years, p<0.0001), were more often diagnosed with septic shock (132 [61%] vs. 134 [43%] patients, p=0.0001), had pre-existent hypertension (141 [65%] vs. 125 [40%] patients, p<0.0001) and CKD (61 [29%] vs. 11 [4%] patients, p<0.0001), received cardiovascular and renal organ support more often (vasopressors: 156 [72%] vs. 164 [53%] patients, p<0.0001; RRT: 37 [17%] vs. 11 [4%] patients, p<0.0001), and had higher disease severity scores (median SOFA score: 9 [IQR 6–11] vs. 6 [IQR 4–8] points, p<0.0001; median APACHE II score: 19 [IQR 14–23] vs. 14 [IQR 10–18] points, p<0.0001) and had a higher mortality rate (28-day mortality: 63 [29%] vs. 44 [14%] patients, p=0.0001; 90-day mortality: 83 [38%] vs. 61 [20%] patients, p<0.0001).

### Diagnosis of acute kidney injury

Acute kidney injury within 48 hours after ICU admission was diagnosed in 234 (44%) patients, with 69 (13%) patients developing stage 1 AKI_48h_, 46 (9%) patients developing stage 2 AKI_48h_ and 119 (23%) patients developing stage 3 AKI_48h_. The median penKid levels at ICU admission were proportionally higher with AKI_48h_ severity (no AKI_48h_: 49.8 [IQR 37.9–75.1] pmol/L, AKI_48h_ stage 1: 111.8 [IQR 68.4–143.2] pmol/L, AKI_48h_ stage 2: 145.8 [IQR 97.5–184.4] pmol/L, AKI_48h_stage 3: 165.1 [111.2–253.3] pmol/L, p<0.0001). Acute kidney injury within 48 hours was early diagnosed by penKid, with an AUC of 0.87 (95% CI 0.84–0.90) (Figure 1, Table 2). Further diagnostic performance characteristics are given in Table 2.

**Figure 1:**
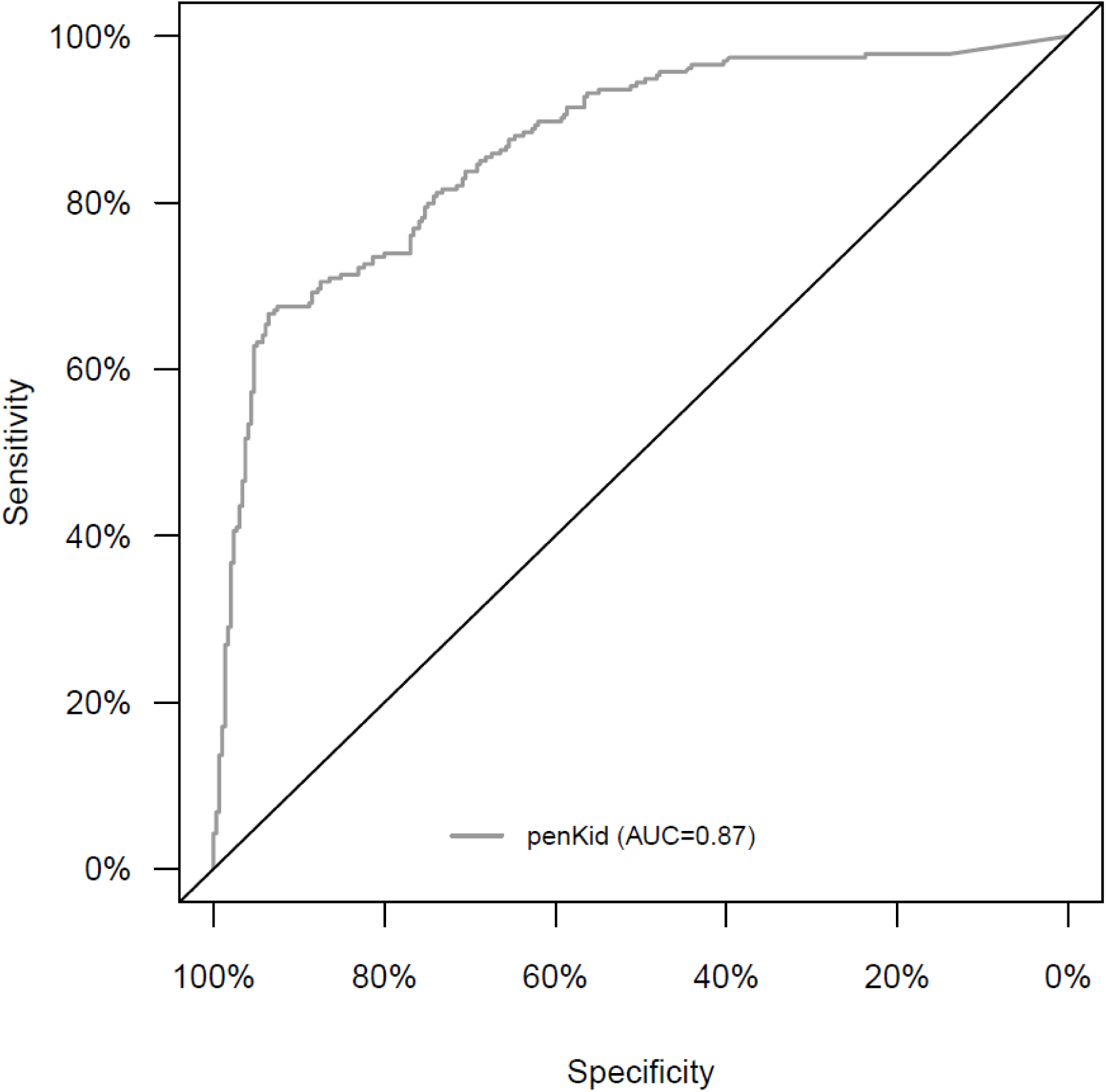
Receiver operating characteristics of proenkephalin A 119–159 for the diagnosis of acute kidney injury. Receiver operating characteristics (ROC) analysis illustrating the performance of proenkephalin A 119–159 (penKid) for the diagnosis of acute kidney injury (AKI) in sepsis or septic shock patients within 48 hours after admission to the intensive care unit. The ROC analysis revealed an area under the curve (AUC) of 0.87 (95% confidence interval [CI] 0.84– 0.90). *AKI, acute kidney injury; AUC, area under the curve; CI, confidence interval; penKid, proenkephalin A 119–159; ROC, receiver operating characteristic*

**Table 2:** Diagnostic performance characteristics of proenkephalin A 119–159 for diagnosis of acute kidney injury. The diagnostic performance characteristics of penKid for diagnosis of acute kidney injury in sepsis or septic shock patients within 48 hours after admission to the intensive care unit were calculated for all patients and subgroups with or without a history of chronic heart failure or hypertension from the AdrenOSS-1 study.

| Patients | AUC | Performance of penKid at cutoff 89 pmol/L |  |  |  |  |  |
| --- | --- | --- | --- | --- | --- | --- | --- |
|  |  | Sensitivity (%) | Specificity (%) | LR+ | LR- | PPV (%) | NPV (%) |
| All patients<br>(n=529;<br>events=234<br>[44%]) | 0.87<br>(0.84–<br>0.90) | 72 (66–<br>77) | 83 (78–<br>87) | 4.2 | 0.34 | 77 (71–<br>82) | 79 (74–<br>83) |
| No CHF<br>(n=467;<br>events=194<br>[42%]) | 0.87<br>(0.84–<br>0.91) | 73 (66–<br>79) | 84 (79–<br>87) | 4.4 | 0.33 | 76 (69–<br>81) | 81 (76–<br>85) |
| CHF<br>(n=46;<br>events=29<br>[63%]) | 0.81<br>(0.70–<br>0.93) | 65 (48–<br>79) | 75 (53–<br>89) | 2.6 | 0.47 | 82 (63–<br>92) | 56 (37–<br>72) |
| No<br>hypertension<br>(n=260;<br>events=80<br>[31%]) | 0.90<br>(0.86–<br>0.94) | 70 (59–<br>79) | 89 (84–<br>93) | 6.6 | 0.34 | 75 (64–<br>83) | 87 (81–<br>91) |
| Hypertension<br>(n=266;<br>events=151<br>[57%]) | 0.82<br>(0.77–<br>0.87) | 73 (65–<br>79) | 73 (64–<br>80) | 2.7 | 0.37 | 78 (71–<br>84) | 67 (59–<br>75) |
| No<br>hypertension<br>and no CHF<br>(n=249;<br>events=74<br>[30%]) | 0.91<br>(0.87–<br>0.95) | 72 (61–<br>81) | 90 (84–<br>93) | 7.0 | 0.32 | 75 (63–<br>83) | 88 (83–<br>92) |

*AKI, acute kidney injury; AUC, area under the curve; CHF, chronic heart failure; LR+, positive likelihood ratio, LR-, negative likelihood ratio; NPV, negative predictive value; penKid, proenkephalin A 119–159; PPV, positive predictive value; number in brackets represent 95% confidence interval*

The AUCs varied between 0.81 (patients with chronic heart failure) and 0.91 (patients without chronic heart failure and hypertension) in the subgroups of patients with chronic heart failure and/or hypertension (Table 2). Lower AUCs were obtained in patients with chronic heart failure and patients with hypertension though these subgroups revealed higher AKI_48h_ rates of 63% and 57%, respectively.

### Decision-making concept using proenkephalin A 119–159

To translate the penKid results into clinical actions, a decision map was designed (Figure 2) based on two penKid cut-offs, resulting in three interpretation bands (Table 3). The penKid cut-offs were derived from augmented sensitivity (>92%; 54 pmol/L) and augmented specificity (>92%; 105 pmol/L). The post-test probability for AKI_48h_ increased from 44% pre-test probability to 87% in patients with a penKid level above 105 pmol/L (LR+ 8.7), allowing a strong rule-in diagnosis of AKI_48h_ in 34% of patients, whereas the post-test probability decreased to 11% in patients with a penKid level below 54 pmol/L (LR-0.15), which was useful for ruling out AKI in 36% of patients. The 28-day mortality rates were impacted accordingly; i.e., for penKid levels below 54 pmol/L, the 28-day mortality was 20% in the total cohort compared to 10%, almost as low as that in non-AKI patients (23 of 268 [9%] non-AKI patients died within 28 days). For penKid levels above 105 pmol/L, the 28-day mortality rate was 30%. The remaining patients with penKid levels ranging from 54 to 105 pmol/L had marginally lower post-test probabilities for AKI (35%) and a slightly higher 28-day mortality rate of 21%.

**Figure 2:**
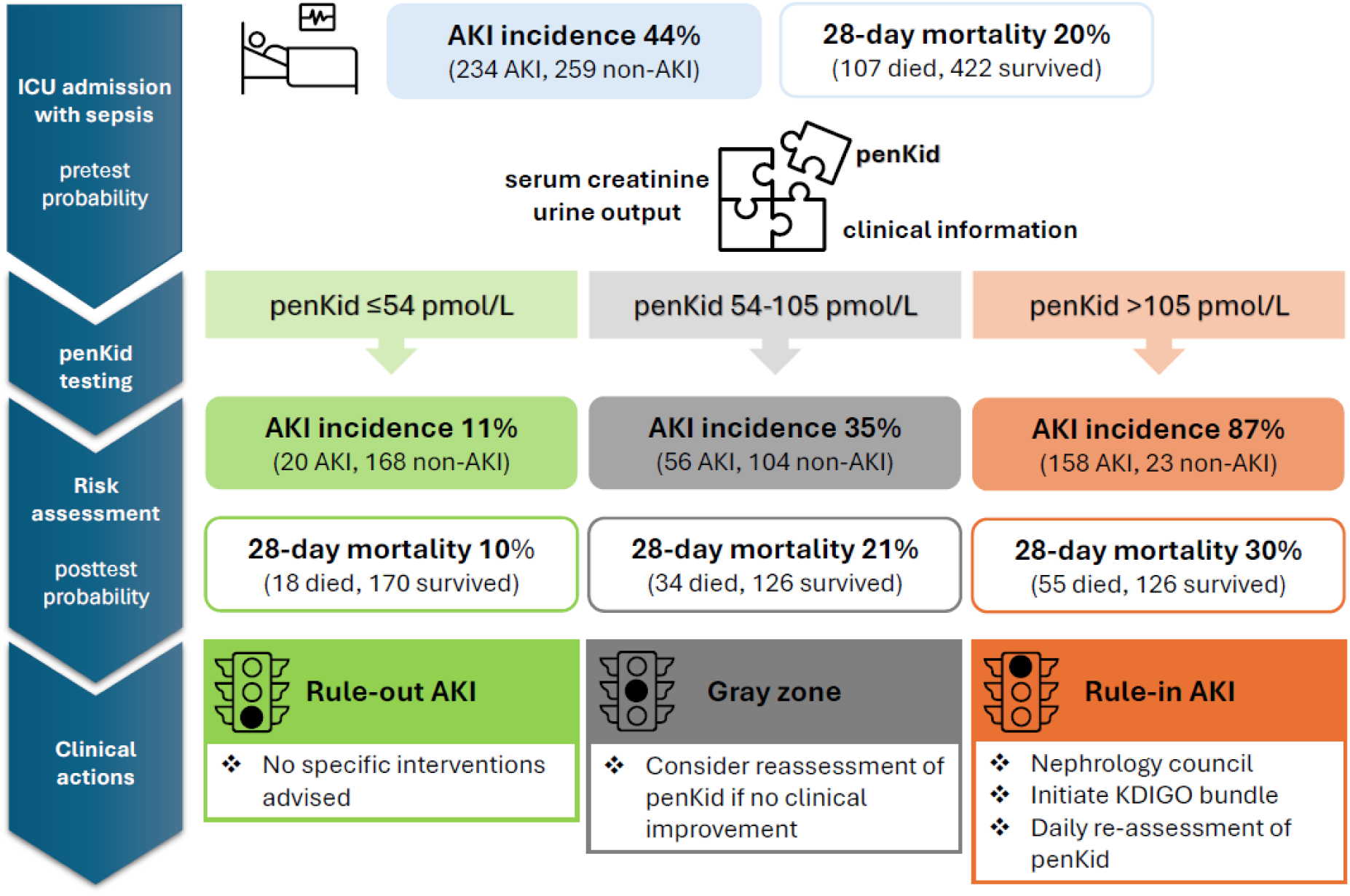
Decision map using proenkephalin A 119–159 as a new kidney function biomarker. Based on the three interpretation bands derived in Table 3, the proenkephalin A 119–159 (penKid) results were translated into clinical actions. These actions are focused on the prevention and management of acute kidney injury (AKI) in addition to the key elements in sepsis therapy (i.e., early and appropriate treatment with antimicrobials, source control, targeted administration of fluids, and vasoactive drugs). For more detailed recommendations, refer to the consensus report of the Acute Disease Quality Initiative group on sepsis-associated AKI (23). *AKI, acute kidney injury; penKid, proenkephalin A 119–159; RRT, renal replacement therapy*

**Table 3:** Acute kidney injury probability based on proenkephalin A 119–159 interpretation bands.

|  |  | No AKI (n) | AKI (n) | Patients in band | Nominal LR | AKI probability | Outer band specificity/sensitivity |
| --- | --- | --- | --- | --- | --- | --- | --- |
| All patients |  | 295 | 234 |  |  | 44% |  |
| penKid range | >105 pmol/L | 23 | 158 | 34% | 8.7 | 87% | 92% rule-in specificity |
|  | 54–105 pmol/L | 104 | 56 | 30% | 0.68 | 35% |  |
|  | ≤54 pmol/L | 168 | 20 | 36% | 0.15 | 11% | 92% rule-out sensitivity |

*AKI, acute kidney injury; LR, likelihood ratio; penKid, proenkephalin A 119–159*

## Discussion

Based on current clinical performance studies, we established the reference range of sphingotest® penKid® in healthy adult subjects, with an adequate representation of sex and age subgroups, and the performance characteristics of sphingotest® penKid® for the early diagnosis of AKI within 48 hours after admission to the ICU in sepsis and septic shock patients, with the upper normal range of 89 pmol/L used as the clinical cutoff.

The penKid data in the reference population are in line with previously published data which reported a 97.5^th^ percentile of 83 pmol/L (95%CI 70–92 pmol/L) in 100 healthy donor samples (24). Reference ranges and diagnostic performance characteristics have not been evaluated in subjects <18 years of age or in pregnant women. The diagnostic performance to detect AKI early in sepsis and septic shock patients supports results from previous studies in patients with suspected or confirmed sepsis or septic shock in the emergency department, ICU, or other clinical settings that used varying penKid cut-off values between 67 pmol/L and 154.5 pmol/L, yielding AUCs ranging from 0.73 to 0.91 (16, 17, 26–29). A meta-analysis, including the aforementioned studies, as well as additional studies in critically ill sepsis and non-sepsis patients, revealed an overall AUC of 0.77 (95%CI 0.73–0.81) (20). In line with the meta-analysis, which reported lower specificity in patients with pre-existent chronic heart failure and hypertension, the current study also revealed decreased specificity of sphingotest® penKid® in septic patients with a history of CHF and/or hypertension, i.e., a higher rate of false-positive results, which may be attributed to an elevated baseline penKid level in these patients relating to the biological role of enkephalins in the cardiovascular system (30). In addition, a diminished overall specificity may have been obtained due to false-positive penKid results in patients that already had AKI at ICU admission but did not further increase in serum creatinine after 48 hours and were therefore grouped as non-AKI in the present analysis. The dilemma of how to determine baseline kidney function in patients presenting as medical emergency with no previous laboratory results on hand is a major drawback of the consensus definition of AKI (12). Furthermore, an AKI definition solely based on serum creatinine and need of RRT may have overlooked AKI cases identified by a reduced urine output resulting in potentially false-positive penKid results. Indeed, there is a frequent underutilization of urine output despite its value for early AKI diagnosis found in a systematic literature review (31). However, AKI severity assessed by urine output revealed a lower mortality risk than AKI staging based on serum creatinine (32).

In addition to early AKI diagnosis, the severity of AKI can be reflected by penKid, i.e., higher penKid levels at ICU admission were observed in patients that were diagnosed with higher AKI stages or the need for RRT in the current study, confirming the proportionality also observed in previous studies (16, 26, 27, 33–37). Considering the strong association between penKid levels and true GFR (15, 38) and the multifaceted nature of sepsis-associated AKI, applying several cut-off values of sphingotest® penKid® in addition to standard-of-care diagnostics may further aid in clinical decision making and targeted management, adapted to AKI probability and 28-day mortality risk. Our analysis recommends the rule-in of ‘likely to develop AKI’ for penKid levels above 105 pmol/L, close monitoring in patients with penKid levels between 54 pmol/L and 105 pmol/L (gray zone) to prevent (further) kidney injury, and the rule-out of ‘unlikely to develop AKI’ for those with penKid levels below 54 pmol/L. Specifically, this allows clear rule-in and rule-out decisions of AKI as early as on ICU admission in two-thirds of sepsis and septic shock patients. Approximately one-third of patients presented with penKid levels between the most actionable rule-in or rule-out interpretation bands, with higher false-positive and false-negative rates, which may be attributed to the large proportion of patients with stage 1 AKI, which are likely to recover within days. This gray zone may also reflect the inaccuracy of the current diagnostic standard and underlines the importance of adding novel biomarkers to traditional parameters to refine AKI staging, as suggested by the Acute Disease Quality Initiative group (23, 39). In fact, disregarding biomarkers that are associated with kidney damage or function and simplification by single cut-off analyses may lead to the loss of important diagnostic information. Retesting sphingotest® penKid® could increase the accuracy of AKI diagnosis and its severity assessment and allow monitoring of the progression or recovery of AKI, and the chance to need therapeutic interventions such as RRT (16). Indeed, serial measurements of penKid have shown added value for the prediction of e.g. AKI and survival rates in sepsis patients (16, 28), and an association between penKid levels measured on day 3 of RRT and successful liberation from RRT was demonstrated in two cohorts of ICU patients (18, 19). Increased biomarker levels indicate kidney injury and renal functional decline before the KDIGO criteria for AKI are met (so-called subclinical AKI, new AKI stage 1S), which helps trigger early diagnostic and (secondary) preventive measures (39, 40). In line with penKid showing a strong correlation with the gold standard method of iohexol or iothalamate clearance for GFR determination, and a penKid-based eGFR formula outperforming existing eGFR formulas (15, 38), subclinical AKI defined by increased penKid was identified in 12% to 18% of septic patients without AKI based on KDIGO criteria and 39% of critically ill patients, and was associated with a risk of death close to that of patients that did suffer from AKI (41, 42). In addition to further clinical assessment and the currently limited routine diagnostic parameters to diagnose AKI, sphingotest® penKid® may contribute to a more timely and accurate diagnosis of AKI and support subsequent treatment decisions, especially in critically ill patients.

Our study has limitations. Due to the retrospective design and nonavailability of data an adapted AKI diagnosis was applied that may have affected AKI prevalence and diagnostic accuracy of sphingotest® penKid®. This study focused on early sepsis-associated AKI in the ICU and the established cut-offs for sphingotest® penKid® may not be generalizable to a broader ICU population or patients from the emergency department. Results need to be confirmed in prospective studies, including septic and nonseptic patients at risk of AKI, and taking potential updates in the AKI definition into account.

## Conclusions

In conclusion, our results from two clinical performance studies showed the added value of sphingotest® penKid® reported in early diagnosis of sepsis-associated acute kidney injury and provided cutoff values focused on actionability in clinical practice.

## Declarations

### Ethics approval and consent to participate

The present study was conducted in France, Belgium, the Netherlands, Italy, and Germany. The study protocol was approved by the local ethics committees, and the study was conducted in accordance with Directive 2001/20/EC as well as good clinical practice (International Conference on Harmonization Harmonized Tripartite Guideline version 4 of May 1, 1996, and decision of November 24, 2006) and the Declaration of Helsinki. Patients were included from June 2015 to May 2016.

### Consent for publication

Not applicable.

### Availability of data and materials

AM and PFL had full access to all the data in the study, and take responsibility for the integrity of the data and the accuracy of the data analysis.

### Authors’ contributions

FD, JS, PFL, PP, and all further investigators of the AdrenOSS-1 study were responsible for the acquisition of data. OH performed the statistical analyses. FU, PP, and PFL contributed to the conception and design of the study. The manuscript was drafted by JS. All the authors were involved in the interpretation of the data and critical revision of the article. The study was sponsored by SphingoTec GmbH and supervised by PFL as principal investigator.

## Data Availability

All data produced in the present work are contained in the manuscript.

## Acknowledgements

Listing of site investigators of the AdrenOSS-1 study

| **Center** | **Investigator** |
| --- | --- |
| Cliniques Universitaires Saint-Luc, Brussels, Belgium | Laterre |
| Clinique St Pierre, Ottignies, Belgium | Dugernier |
| Hôpital Jolimont, Haine-St-Paul, Belgium | Huberlant |
| Klinik für Operative Intensivmedizin und Intermediate Care, Universitätsklinikum der RWTH, Aachen, Germany | Marx |
| Klinik für Anästhesiologie und Operative Intensivmedizin, Universitätsklinikum Köln, Köln, Germany | Hohn |
| HELIOS-Klinikum Erfurt, Erfurt, Germany | Meier-Hellmann |
| Klinikum Augsburg, Augsburg, Germany | Jaschinski |
| Klinik für Anästhesiologie und Intensivmedizin, Jena, Germany | Kortgen |
| CHU Dupuytren, Limoges, France | Francois |
| CHD les Oudairies, La Roche sur Yon, France | Lascarrou |
| CHU de Tours, Tours, France | Mercier |
| Centre hospitalier d'Angoulême, Angoulême, France | Desachy |
| CHU Angers, Angers, France | Lasocki |
| Hôpital Lariboisière, Paris, France (two centers) | Mebazaa |
| Hôpital Saint-Louis 1, Paris, France | Jacob |
| Hôpital Louis Mourier, Colombes, France | Gaudry |
| Hôpital Hautepierre, Strasbourg, France | Pottecher |
| CHU Estaing, Clermont Ferrand, France | Constantin |
| Hôpital Bichat Claude-Bernard, Paris, France | Sonneville |
| Sant'Andrea Hospital, Rome, Italy | Di Somma |
| Policlinico Universitario A. Gemelli, Rome, Italy | Antonelli |
| Medisch Spectrum Twente, Enschede, The Netherlands | Beishuizen |
| UMC Radboudziekenhuis, Nijmegen, The Netherlands | Pickkers |

## Principal investigator

Prof. Dr. Pierre-François Laterre, Head of Department of Intensive Care Medicine, CHR Mons-Hainaut, Mons, Belgium, phone: + 32 2764 27 35, fax: + 32 2764 89 28

## List of abbreviations

AdrenOSS-1: Adrenomedullin and Outcome in Sepsis and Septic Shock-1
AKI: acute kidney injury
APACHE II: acute and physiology and chronic health evaluation II
AUC: area under the curve
CI: confidence interval
CKD: chronic kidney disease
CLSI: Clinical and Laboratory Standards Institute
EDTA: ethylenediaminetetraacetic acid
GFR: glomerular filtration rate
ICU: intensive care unit
ILMA: immunoluminometric assay
IQR: interquartile range
KDIGO: Kidney Disease: Improving Global Outcomes
LR+: positive likelihood ratio
LR-: negative likelihood ratio
MTP: microtiter plate
NPV: negative predictive value
OR: odds ratio
penKid: proenkephalin A 119–159
PPV: positive predictive value
ROC: receiver operating characteristics
RRT: renal replacement therapy
SOFA: sequential organ failure assessment

